# Changing patterns of mortality during the COVID-19 pandemic: population-based modelling to understand palliative care implications

**DOI:** 10.1101/2020.06.07.20124693

**Authors:** Anna E Bone, Anne M Finucane, Javiera Leniz, Irene J Higginson, Katherine E Sleeman

## Abstract

**Background:** COVID-19 has directly and indirectly caused high mortality worldwide.

**Aim:** To explore patterns of mortality during the COVID-19 pandemic and implications for palliative care provision, planning, and research.

**Design:** Descriptive analysis and population-based modelling of routine data.

**Participants and setting:** All deaths registered in England and Wales between 7^th^ March and 15^th^ May 2020. We described the following mortality categories by age, gender and place of death: 1) baseline deaths (deaths that would typically occur in a given period) 2) COVID-19 deaths 3) additional deaths not directly attributed to COVID-19. We estimated the proportion of COVID-19 deaths among people who would be in their last year of life in the absence of the pandemic, using simple modelling with explicit assumptions.

**Results:** During the first 10 weeks of the pandemic there were 101,615 baseline deaths, 41,105 COVID-19 deaths and 14,520 additional deaths. Deaths in care homes increased by 220% compared to home and hospital deaths which increased by 77% and 90%. Hospice deaths fell by 20%. Additional deaths were among older people (86% aged ≥75 years), and most occurred in care homes (56%) and at home (43%). We estimate that 44% (38% to 50%) of COVID-19 deaths occurred among people who would have been in their last year of life in the absence of the pandemic.

**Conclusions:** Healthcare systems must ensure availability of palliative care to support people with severe COVID-19 in community and hospital settings. Integrated models of palliative care in care homes are urgently needed.

**Key statements:** *What is already known about the topic?:*

- The COVID-19 pandemic has directly and indirectly resulted in high mortality in many affected nations.
- Internationally the response has been focused on prevention and curative treatments, with little emphasis on palliative care needs of people dying during the COVID-19 pandemic.
- We do not know how many of those dying with COVID-19 would have been in their last year of life in the absence of the pandemic, and this group may have distinct care needs.

*What this paper adds:*

- The number of people dying in care homes trebled during the first 10 weeks of the COVID-19 pandemic in England and Wales; many of these deaths were ‘additional deaths’, that is associated with the COVID-19 pandemic but not directly as a result of COVID-19.
- We estimate almost half of all COVID-19 deaths occurred among people who would have been in their last year of life in the absence of the pandemic.

*Implications for practice, theory or policy:*

- Healthcare systems must ensure availability of palliative care to support people with severe COVID-19 in community and hospital settings.
- The need for integrated models of palliative care in care home settings is imperative and research to underpin these models is warranted.

## Introduction

Approaches to death and dying reveal much of the attitude of society as a whole to the individuals who compose it (Cicely Saunders, 1984).^1^ The COVID-19 pandemic has led to an unprecedented surge in mortality around the world, with some countries such as United Kingdom (UK), the United States, and Italy particularly affected. In the UK and elsewhere the government response has been focused on prevention and curative treatments. There has been very little emphasis internationally on care needs, including palliative care needs, of people dying during the COVID-19 pandemic.^2^

Clinical research and observation has found that people dying with COVID-19 are symptomatic, frequently experiencing breathlessness and agitation near the end of life.^3^ Clinical uncertainty around the illness trajectories of those infected increases the complexity of clinical decision-making and communication with patients and family.^4^ Deaths occurring during the COVID-19 pandemic are likely to be associated with poorer bereavement outcomes for family and friends and higher distress and support needs among frontline health and social care staff.^5^ There is an important role for palliative care but these needs have not been described at a population level.

Large scale population-based studies exploring patterns of mortality are important for quantifying numbers in need of care, and for informing governments about the need to plan and reorganise services. In England and Wales, official mortality data from the Office for National Statistics (ONS) reveal that during the first 10 weeks of the pandemic, (7^th^ March to 15th May), there were over 41,000 COVID-19 deaths, most occurring in hospital (65%), with 28% in care homes and few elsewhere (7%).^6^ The ONS estimated that there were also over 14,000 ‘additional’ deaths which are deaths over and above the number of deaths expected based on the 5-year average, that were not certified as caused by COVID-19.^6^

This study aims to explore patterns of mortality during the first 10 weeks of the COVID-19 pandemic in England and Wales (7^th^ March to 15^th^ May 2020) to understand implications for palliative care. The objectives are: 1) to explore trends in place of death; 2) to explore the age and gender distribution of baseline deaths, COVID-19 deaths and additional deaths; 3) to estimate the proportion of people who died from COVID-19 who would have been in their last year of life, and differences by age; 4) to use this information to discuss implications for palliative care provision, service planning, and research.

## Methods

### Design and setting

Descriptive analysis and population-based modelling using routinely collected mortality data for England and Wales.

### Data

We used publicly available data from the Office for National Statistics including:

- Weekly registered deaths in England and Wales by age group for 2015 to 2019.
- Weekly registered deaths in England and Wales in the weeks corresponding to the COVID-19 pandemic, between 7^th^ March (week 11) and 15^th^ May 2020 (week 20)^6^. This includes all deaths and COVID-19 deaths by age, gender and place of occurrence.
- Principal population projections of the total population of England and Wales by age for 2020 (2018-based)^7^

### Analysis

We first defined the following mortality categories: baseline deaths, COVID-19 deaths and additional deaths, as detailed in Box 1.

**Box 1. Definitions of mortality categories**

| Terms | Definition |
| --- | --- |
| **Baseline deaths** | Deaths that would be expected to occur in a typical year without a pandemic. For this analysis we used the average number of deaths that occurred in the same period over the past five years. |
| **COVID-19 deaths** | Deaths officially registered as with or from COVID-19. This category includes a subgroup: |
| **a. COVID-19 deaths among annual baseline deaths** | a. COVID-19 deaths among those who would have been in their last year of life in the absence of the pandemic. |
| **Additional deaths** | Deaths in excess of baseline deaths that are not accounted for by deaths certified as being as a direct result of COVID-19. This group might include deaths due to COVID-19 but not recorded as such, or deaths indirectly related to the pandemic for example through avoidance of hospital care. |

The numbers of people in each mortality category were calculated for England and Wales between 7^th^ March and 15^th^ May 2020 as follows:

#### 1. Baseline deaths

We estimated the baseline deaths according to the average deaths in the 10-week period of interest (weeks 11 to 20) over the years 2015 to 2019, by age and gender.

#### 2. COVID-19 deaths

Data on COVID-19 deaths was obtained from the ONS weekly registered death, which includes deaths by age, gender and place of occurrence.^6^ The ONS produce figures for deaths involving COVID-19, based on any mention of COVID-19 on the death certificate, including where the physician suspects COVID-19 based on symptoms but where no test was available.^6^ These figures include deaths outside of hospital.

##### a). COVID-19 deaths among annual baseline deaths

First, we derived an estimate of the proportion of the population infected in each age group, by re-arranging the following equation:

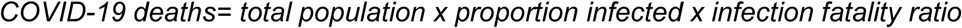

The information we used in this equation included the number of registered COVID-19 deaths reported by ONS^6^; the total population from the ONS principal projections^7^; and infection fatality ratios by 10-year age groupings obtained from Ferguson et al.^8 9^(supplementary table 1).

**Table 1.** COVID-19 deaths over 10-week period from 7^th^ March to 15^th^ May 2020 among annual baseline deaths

| Age | Total population of England and Wales<sup>7</sup> | Baseline deaths over 12 months (based on 5 year average) | Total COVID-19 deaths up to 15<sup>st</sup> May | Infection fatality ratio (based on Ferguson et al 2020)<sup>8</sup> | Estimated proportion infected with COVID-19 over the period | COVID-19 deaths among annual baseline deaths n (% of all COVID-19 deaths) |  |  |
| --- | --- | --- | --- | --- | --- | --- | --- | --- |
|  |  |  |  |  |  | Multiplier of 7 -primary estimate | Multiplier of 6 - lower estimate | Multiplier of 8- upper estimate |
| 0 to 9 | 7,142,647 | 3,305 | 3 | 0.00% | 2.1% | 0 (0%) | 0 (0%) | 0 (0%) |
| 10 to 19 | 6,908,476 | 2,823 | 10 | 0.01% | 2.4% | 0 (0%) | 0 (0%) | 0 (0%) |
| 20 to 29 | 7,654,985 | 4,955 | 64 | 0.03% | 2.8% | 0 (0%) | 0 (0%) | 0 (0%) |
| 30 to 39 | 7,973,235 | 4,955 | 181 | 0.08% | 2.8% | 1(0%) | 1(0%) | 1(0%) |
| 40 to 49 | 7,495,052 | 18,176 | 591 | 0.15% | 4.3% | 10 (2%) | 9(1%) | 11 (2%) |
| 50 to 59 | 8,080,681 | 31,396 | 1,953 | 0.6% | 4.0% | 57 (3%) | 46(2%) | 61 (3%) |
| 60 to 69 | 6,371,654 | 59,543 | 4,150 | 2.2% | 3.0% | 271 (7%) | 233 (6%) | 310 (7%) |
| 70 to 79 | 5,149,885 | 119,014 | 9,431 | 5.1% | 3.6% | 1,526 (16%) | 1,308 (14%) | 1,744 (18%) |
| 80+ | 3,052,619 | 286,883 | 24,722 | 9.3% | 8.7% | 16,264 (66%) | 13,940 (56%) | 18,587 (75%) |
| Total | 59,829,234 | 531,050 | 41,105 | - | - | 18,125 (44%) | 15,536 (38%) | 20,714 (50%) |

Second, we hypothesised that someone who is in their last year of life will have an increased risk of dying from COVID-19 compared to someone in the same age group who is not in their last year of life. To account for this, we applied a risk of dying multiplier to this group based on evidence of increased risk of death among people with serious conditions (supplementary Table 2).^10^ We applied a range of multipliers, with our primary estimate based on the median risk of death across different conditions.^10^ Therefore, the number of people who die with COVID-19 who would be expected to die within a year was calculated using the following equation:

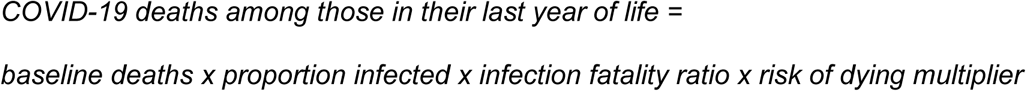

#### 3. Additional deaths

We calculated additional deaths by subtracting the estimated baseline deaths and COVID- 19 deaths from all registered deaths on a per week basis over the 10-week period of interest (weeks 11 to 20). This was calculated for each age and gender group separately to identify additional deaths within different demographic categories. Where this resulted in a negative value, the estimated baseline figure was reduced by this amount and the value for additional deaths was zero.

#### Age, gender and place of death

To understand the age and gender distribution of the COVID-19 and additional deaths we grouped data into 4 age groups (<45, 45-64, 65-74, and ≥85 years) and compared them with baseline deaths over the period of interest (weeks 11 to 20). We described trends in place of occurrence of deaths, using the ONS defined categories of ‘hospital’ (acute and community, not psychiatric), ‘care home’ (nursing and residential), ‘home’, ‘hospice’ and combined ‘other communal’ and ‘elsewhere’. To estimate additional deaths by place of death over the period, we used place of death data for week 11 to estimate baseline place of death, as weekly place of death information is unavailable for previous years.

### Patient and public involvement

We sought the views and experiences of members of our patient and public network regarding their concerns around COVID-19 pandemic (Johnson et al, 2020 submitted).^11^ These consultations informed the focus and design of this study. A representative from the network provided feedback on the manuscript.

### Ethics

This study uses routinely collected anonymised and publicly available data. No ethical approvals were necessary.

## Results

Between 7^th^ March and 15^th^ May 2020 (weeks 11 to 20) in England and Wales there were 157,239 deaths of which 41,105 were from COVID-19. We estimate that this included 101,615 baseline deaths (as estimated by average weekly deaths over the same period during 2015-2019), and 14,520 additional deaths (Figure 1).

**Figure 1.**
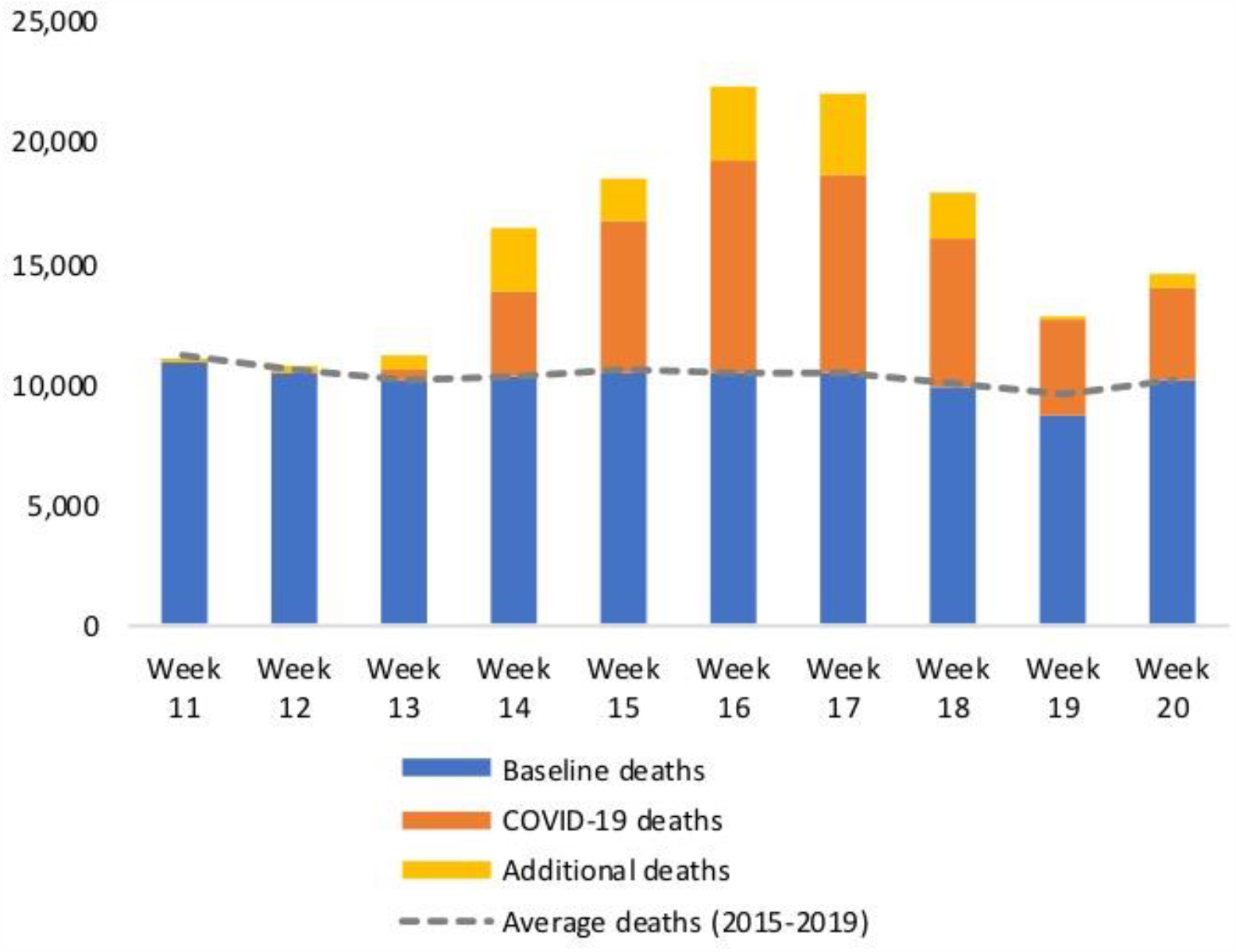
All registered deaths during the COVID-19 pandemic between 7th March and 15th May 2020 (weeks 11 to 20) in England and Wales.

### Place of death during the pandemic

Where people die in England and Wales has changed (Figure 2). Taking all deaths together, weekly hospital deaths increased by 90% from week 11 (n=4,975) to a peak in week 16 (n=9,434). We observe a 77% increase in home deaths between weeks 11 (n=2,725) and 17 (n=4834), while care home deaths increased by 220% during the same period (week 11 n=2,471, week 17 n= 7,911). In week 18, care homes temporarily became the most common place to die. Hospice deaths fell by 20% between weeks 11 (n=553) and week 19 (n=441), representing 3% of deaths during the 10 week period.

**Figure 2.**
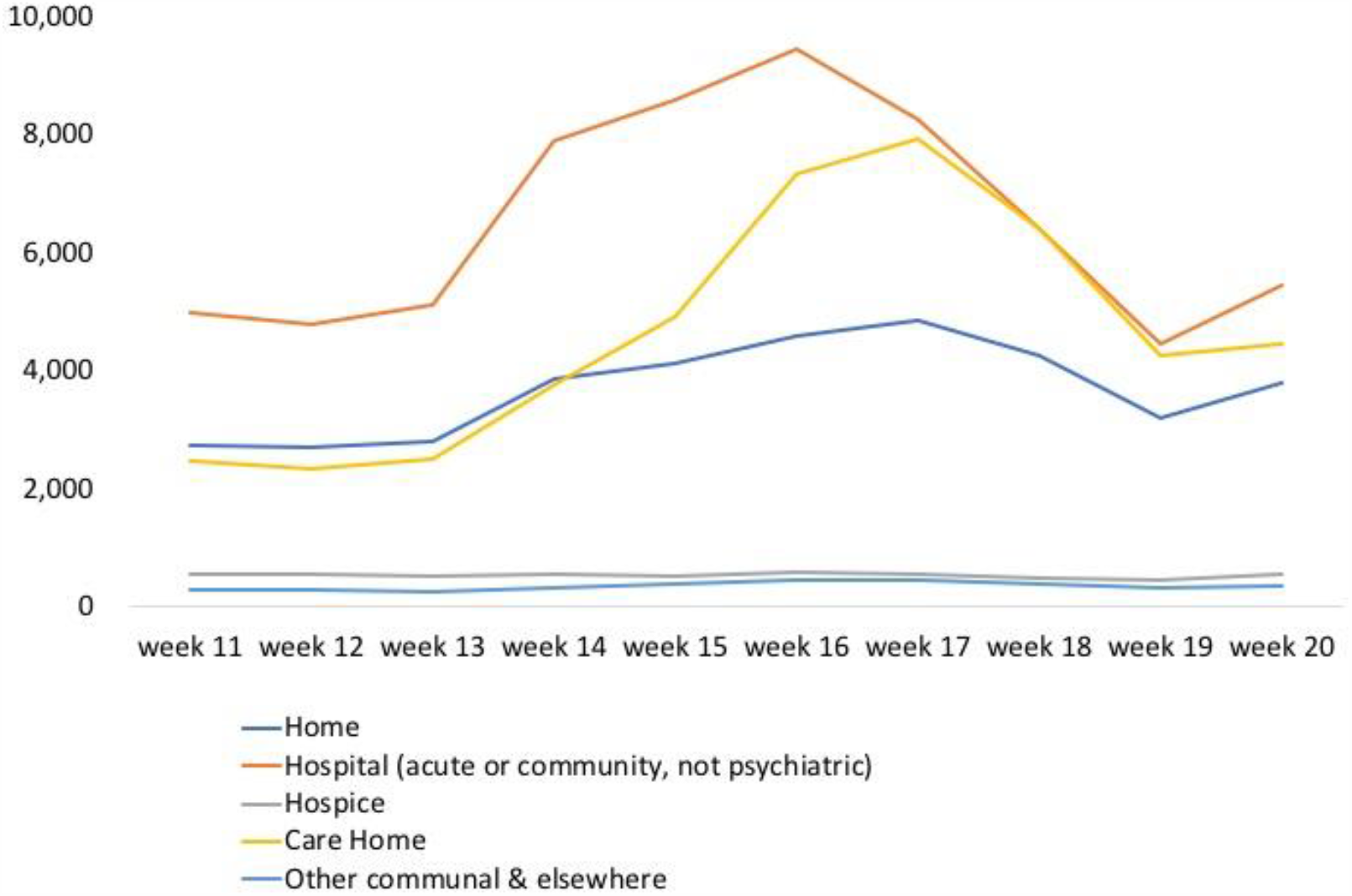
All registered deaths during the COVID-19 pandemic between 7th March and 15th May 2020 (weeks 11 to 20) in England and Wales by place of occurrence.

Most COVID-19 deaths occurred in hospital (65%) and care homes (28%) with few occurring at home and hospice (5% and 1%). The location of the additional deaths has not been described. Comparing place of death for additional deaths occurring in weeks 12-20 with week 11, we found the majority of additional deaths occurred in care homes (56%) and at home (43%) (Figure 3). During weeks 12-20 there were fewer additional deaths in hospital and hospice compared to week 11, indicating that there were fewer non-COVID-19 deaths in these settings than usual.

**Figure 3.**
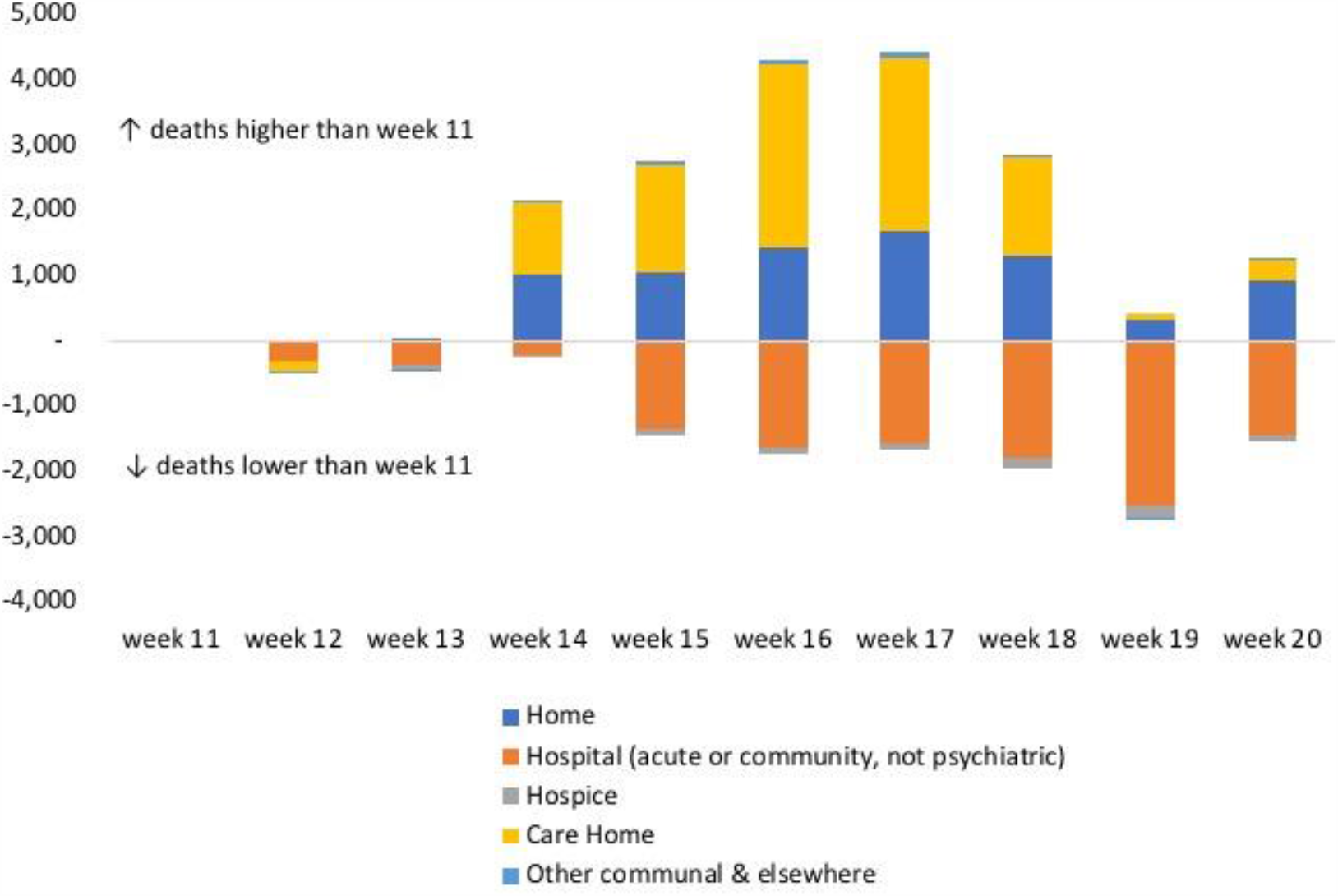
Difference in number of additional deaths (not attributed to COVID-19) in weeks 12 to 20 compared to week 11 in 2020, by place of occurrence.

### Mortality categories by age and gender

We found the age distribution of COVID-19 deaths to be overall older compared to baseline deaths, with 74% of COVID-19 deaths aged ≥75 years compared to 68% of baseline deaths (Figure 4). Additional deaths are older still, with 83% aged ≥75 years and 56% aged ≥85 years. A greater proportion of COVID-19 deaths are male (56%), compared to 49% of baseline deaths and 50% of additional deaths.

**Figure 4.**
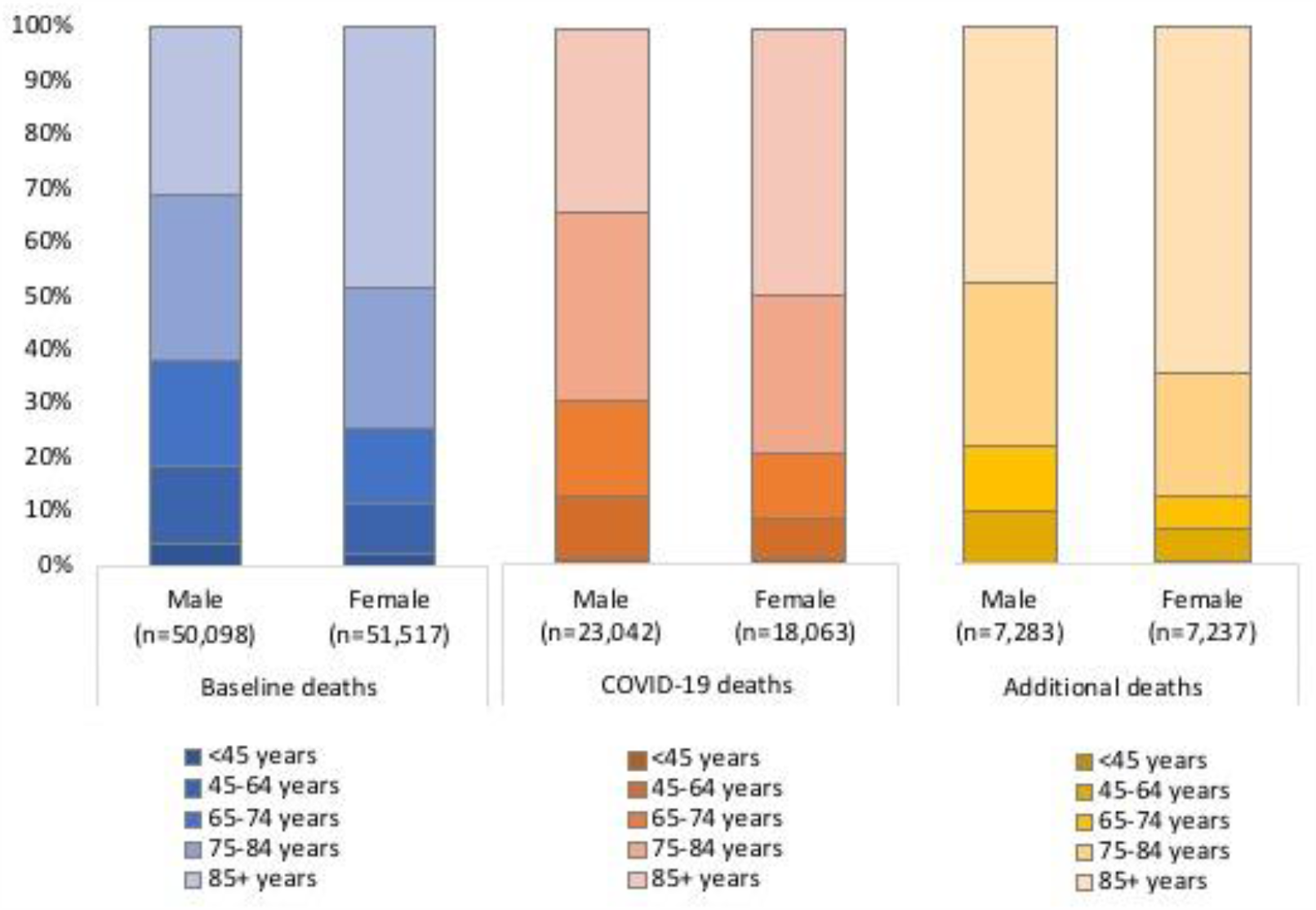
Age and gender distribution of baseline deaths, COVID-19 deaths, and additional deaths between 7th March and 15th May 2020 (weeks 11 to 20) in England and Wales.

### COVID-19 deaths among annual baseline deaths

Among the people who are registered as dying with COVID-19 we have estimated a group who would be in their last year of life in the absence of the pandemic. Using our primary (and lower and upper) estimates for the risk of dying multiplier, we found that 44% (38-50%) of people who died with COVID-19 over the period of interest were likely to be in their last year of life (Table 1 and supplementary figure 1). This increases with age, such that by >80 years, 66% (56-75%) of people who die with COVID are likely in their last year of life in the absence of the pandemic.

## Discussion

### Main findings

Using routine data and modelling scenarios to understand mortality patterns during the COVID-19 pandemic, we highlight that care homes temporarily became the most common place to die in England and Wales, and that hospital and home deaths increased by over 50% while deaths in hospices fell by 20%. We show that people in the additional deaths category are proportionately more likely to be older, and to die in care homes and at home, than those who are certified as dying with COVID-19. Finally, we estimate for the first time that almost half of COVID-19 deaths occur among people who would be in their last year of life in the absence of the pandemic, and that this increases to two thirds of those aged over 85 years.

### Implications for palliative care

Deaths in hospitals, care homes and at home increased during the first weeks of the COVID- 19 pandemic. Rapid adaptation of models of palliative care is needed to support the increased number of people dying in these settings. Guidance on remote consultations in primary care^12^ and on managing distressing symptoms in the community^13^ are available to help support care for people with suspected COVID-19, but challenges remain in terms of workforce and equipment. Specialist palliative care provision in care homes is currently variable,^14-16^ and guidance to support palliative care for people with COVID-19 in nursing homes has been found to be lacking and limited in focus.^17^ In contrast, deaths in hospices fell during the time period. A similar phenomenon was found during the 2003 SARS pandemic and highlighted the need for seamless continuity of care between settings.^18^ Systems that enable hospice resources to be shifted from inpatient to community settings are likely to be important in future pandemic peaks.^19^

Little has been described about the characteristics of the additional deaths that have occurred during the COVID-19 pandemic. The aetiology of these deaths is not known, in particular whether they are undiagnosed COVID-19 deaths, or deaths associated with the pandemic without being as a result of the disease (for example due to avoidance of urgent healthcare).^20^ There is emerging evidence that older people with COVID-19 may present with non-specific and atypical symptoms such as delirium or low-grade fever.^21^ We found that the additional deaths group comprises predominantly older people in care homes, suggesting that these may be undiagnosed COVID-19 deaths. Irrespective of aetiology, this group are likely to have significant palliative care needs. The characteristics of the additional deaths population is likely to change as the pandemic progresses and deferred harm from missed routine check-ups or screening becomes more likely.^22^

At a population level we estimate that 44% of COVID-19 deaths occur among people who would have been in their last year of life in the absence of the pandemic, and that among those aged >85 years 66% are expected to be in their last year of life. For those with advanced conditions living at home and in care homes, guidelines recommend that general practitioners (GPs), community healthcare staff and community geriatricians urgently review advance care plans with patients, and discuss their care preferences in the case of contracting COVID-19.^13, 23^ Our analysis indicates that this group is potentially very large and that enacting this recommendation will take significant resource.

While the focus of this paper is COVID-19, it should be noted that baseline deaths outnumber COVID-19 and additional deaths. Previous modelling has shown that around 69- 82% of the baseline group are likely to have palliative care needs.^24^ COVID-19 will impact on this group for example through fewer face to face contacts, and potentially less availability of specialist palliative care support as services are suddenly stretched beyond their usual bounds.

Across all mortality categories there is the need for bereavement support as a result of higher prevalence of complicated grief.^25^ There will be the typical grief associated with death and dying, but on a much larger scale. In addition there will also be novel grief processes, linked to social isolation and distancing, self-blame around infection, and not being able to attend funerals.^26^ Palliative care specialists with particular expertise in bereavement support have a role in supporting families in coping with bereavement, as well as and supporting colleagues to deliver this care alongside other relevant organisations.

### Strengths and limitations

This is the first paper to estimate and characterise population-level patterns of mortality during the COVID-19 pandemic in relation to palliative care. We have produced simple models with explicit assumptions to raise questions and stimulate discussion. We calculated additional deaths for each age and gender group separately to improve precision. However, this study has limitations. We have been limited by the available official data, including the lack of historical data on place of death by week in England and Wales. We have used data for England and Wales, and patterns are likely to vary by country and region, particularly with respect to care home deaths. The infection fatality ratio and risk of dying multipliers for those in their last year of life were based on latest available data, and this might change over the course of the pandemic.

This study has focused on mortality only. Other aspects of palliative care that are important to consider, for example symptom control, support with difficult conversations, advance care planning, and complex clinical decision-making.^27^ There are also longer-term needs among those surviving COVID-19, such as rehabilitation and palliative care, which are likely to be significant and important for service planning but were not modelled here. We have produced population-based models that are useful for strategic health service planning but are not directly applicable to clinical decision-making at an individual level. For example, while we have estimated the number of people dying from COVID-19 who would have been in their last year of life in the absence of the pandemic at the population level, we acknowledge the challenges of prognostication at the individual level.^28^

### Implications for policy, data and future research

A population level understanding of the palliative care needs of people with severe COVID- 19 is essential to guide efficient health service planning, but has been missing from political rhetoric around the world, much of which has been focused on intensive care capacity. COVID-19 is now the focus of much research but very few studies are focused on palliative and end of life care.^29, 30^ Data and research are urgently required to answer the following questions:

- What are the best integrated models of palliative care in care homes, that can be adapted for use during the COVID-19 pandemic?
- What is the impact of COVID-19 on the hospice sector and how might hospice services best use their available resources?
- What is the impact on the baseline group of the additional demand for palliative care?
- What is the symptom burden among people dying with COVID-19 in different settings?
- What is the impact of COVID-19 on complex grief and what resources are needed to support those experiencing this?
- What is the impact of COVID-19 on carers supporting people with palliative care needs in community settings?
- What will be the effect of COVID-19 on medium and long-term mortality projections and palliative care need?

## Conclusions

Our study has highlighted the high level of need for palliative care during the COVID-19 pandemic. Healthcare systems responding to COVID-19 must consider the need for palliative care surge capacity, including the workforce and equipment to enable people to be cared for appropriately in all settings. The need for integrated models of palliative care in care home settings is imperative. The rapid changes to practice during the pandemic provide important opportunities for research, evaluation and learning and it is essential that new models are underpinned by evidence. We show that a high proportion of people who die with COVID-19 would be in their last year of life in the absence of the pandemic. While these data are estimates, and underlying assumptions may change, they highlight the need for careful consideration of the palliative care needs, in addition to intensive care needs, of people with severe COVID-19.

## Data Availability

All data is publicly available.

https://www.ons.gov.uk/peoplepopulationandcommunity/birthsdeathsandmarriages/deaths/datasets/weeklyprovisionalfiguresondeathsregisteredinenglandandwales

## Statements

### Contributors

All authors conceived the idea for the study. AEB, AMF and JL conducted the analyses. AEB and KES wrote the first draft of the manuscript. All authors critically revised the manuscript for intellectual content and approved the final version of the submitted manuscript. The corresponding author attests that all listed authors meet authorship criteria and that no others meeting the criteria have been omitted.

## Acknowledgement

We are grateful for the input from Janice Tausig

## Data sharing

All data is publicly available

## Ethical approval

Not required

